# Antenatal care utilisation in Nigeria: assessing disparities between rural and urban areas—analysis of the 2018 Nigeria demographic and health survey

**DOI:** 10.1101/2024.01.24.24301729

**Authors:** Emmanuel O Adewuyi, Asa Auta, Mary I Adewuyi, Aaron Akpu Phili, Victory Olutuase, Yun Zhao, Vishnu Khanal

**Author notes:** Corresponding authors: Emmanuel Adewuyi.

## Abstract

**Objectives:** This study presents a comprehensive assessment of antenatal care (ANC) utilisation in Nigeria, focusing on the disparities between rural and urban areas.

**Methods:** We used the data disaggregation approach to analyse the 2018 Nigeria Demographic and Health Survey. We estimated ANC utilisation, assessed the receipt of ANC components, and identified factors associated with eight or more (≥ 8) ANC contacts nationally and across rural and urban residences.

**Results:** The overall ≥ 8 ANC utilisation was 20.3% in Nigeria—35.5% in urban and 10.4% in rural areas. Nationally and in urban areas, the North-East region had the lowest ANC use at 3.7% and 3.0%, respectively, while the North-West had the lowest in rural areas (2.7%). Nationally, 69% of mothers received iron supplements, 70% had tetanus injections, and 16% received drugs for intestinal parasites, with urban residents having higher percentages across all ANC components. Maternal and husband education, health insurance, and maternal autonomy were common factors associated with increased ANC odds at the national, rural, and urban residences. All ethnic groups had higher ANC odds than the Hausa/Fulanis in urban areas, while only the Yorubas had greater odds in rural areas. Internet use was significant only in the national context, watching television only in urban settings, while maternal working status, wealth, birth type, religion, and listening to the radio were significant only in rural areas.

**Conclusion:** Our study highlights considerable disparities in ANC utilisation and quality with a greater vulnerability for rural residents, rural northern regions, and socioeconomically disadvantaged mothers. Targeted interventions are imperative to address the disparities and improve ANC use in Nigeria, with priority for the most vulnerable sub-populations.

## Background

Antenatal care (ANC) is a vital component of comprehensive maternal healthcare services provided by appropriately skilled health professionals to pregnant women (and adolescent girls) to optimise their health and promote positive pregnancy outcomes for mothers and newborns [1, 2]. These services encompass a range of healthcare assessments, screening for potential risk factors, vaccinations, nutritional supplementation (such as folic acid and iron)[3, 4], prevention or management of pregnancy-related or comorbid disorders, and health promotional activities [1, 2]. By providing a dual focus on public health and clinical intervention, ANC serves as a crucial platform for improving outcomes during pregnancy with the overarching goal of decreasing maternal and perinatal morbidity and mortality [1]. Early detection and treatment of pregnancy-associated disorders, as well as the identification of women exhibiting symptoms suggestive of potential obstetric complications for individualised specialist care, are integral components of ANC [1]. Notably, women with adequate ANC contacts are more likely to deliver their babies in healthcare facilities [5, 6] with opportunities for better access to comprehensive emergency obstetric care services when needed [3, 4]. ANC, thus, plays a pivotal role in creating a continuum of care that addresses the healthcare needs of pregnant women and establishes a foundation for improved maternal and neonatal health outcomes.

In its recent guidelines, in 2016, the World Health Organisation (WHO) recommended a holistic approach to ANC that emphasises woman-centred care tailored to individual needs and prioritises pregnant women’s physical, emotional, and social well-being [1, 2]. Unlike the basic or focused ANC model, which requires a minimum of four visits, the WHO now recommends a minimum of eight ANC contacts throughout the pregnancy, starting early in the first trimester and comprising one contact in the first trimester, two in the second trimester, and five in the third trimester [1, 7]. This recommendation covers various aspects of healthcare service provision, including the necessity for routine medical check-ups, counselling on nutrition, exercise, and childbirth preparation, as well as addressing social determinants of health [1–3]. Moreover, the WHO highlights the importance of continuous support from healthcare providers, respectful communication, and involvement of women in decision-making processes towards enhancing the overall quality of ANC and contributing to positive pregnancy experiences for expectant mothers and their developing foetuses [1–3]. Despite its well-established significance and the WHO’s evidence-based recommendations, ANC remains underutilised in several low-to-middle-income countries, including Nigeria, highlighting the urgent need for concerted efforts in this respect.

Notably, Nigeria bears a disproportionately high burden of global maternal and neonatal mortalities, accounting for over 28% of maternal deaths (> 82,000 in 2020) and ranking second in the absolute number of neonatal deaths with over 270,000 reported deaths in 2019 [8, 9]. These mortalities are substantial and concerning, especially compared to Nigeria’s population, less than 3% of the world’s total [8]. Amidst these alarming statistics, the pivotal role of ANC use becomes evident. Optimal ANC utilisation offers a multifaceted approach to addressing critical health challenges of pregnancy through early detection of diseases or risk factors, implementing preventative measures, offering education and counselling, providing nutritional support, and facilitating access to specialised care. Consequently, efforts focused on enhancing the utilisation of optimal ANC services represent a key strategy in mitigating Nigeria’s high maternal and neonatal mortality burden—aligning with global health objectives [10, 11]. Consistent with this premise, some studies have explored factors associated with ANC service utilisation in Nigeria [12–22]. However, several of these and related studies are limited by their generalised approach, primarily focusing on national estimates, which may inadvertently mask differences between and within population groups. Nigeria has a wide range of demographics and healthcare infrastructure, highlighting the significance of understanding ANC utilisation across various subpopulations [23]. It is essential to design interventions that address the specific challenges faced by different groups within the country to achieve meaningful progress in maternal and neonatal health outcomes, hence, the focus of our study.

In a previous study, we used a data disaggregation approach to gain insight into Nigeria’s under-utilisation of ANC services [13]. Recognising the importance of this subject and the new WHO recommendations, we aim to further our previous work by investigating the utilisation of ANC services in Nigeria, focusing on the disparities between rural and urban areas. We used the latest nationally representative demographic and health survey data in the present study [23]. Moreover, we expanded our investigation to advance knowledge of the disparities in the receipt of essential components of ANC services across rural and urban residences in the country. ANC components include iron supplementation, tetanus vaccinations, interventions for intestinal parasites, blood pressure checks, and various laboratory tests [1, 2]. The components are crucial for promoting maternal and foetal health by detecting and managing potential risks, ensuring optimal nutrition, and providing preventive measures, ultimately contributing to a safer pregnancy and childbirth [1, 2]. The current study, thus, offers evidence-based knowledge that can inform policies towards improving Nigeria’s maternal and neonatal care outcomes. Aligning with global maternal-child healthcare initiatives, our findings are expected to contribute to realising the Sustainable Development Goal (SDG) 3 targets of reducing maternal mortality and neonatal mortality burdens [11] in Nigeria.

## Methods

### Data Source

This study relies on the data extracted from the 2018 Nigerian Demographic and Health Survey (NDHS), a nationwide survey conducted every five years in Nigeria, starting from 1990 [23]. The survey adheres to a validated methodology and is collaboratively implemented by the Nigerian National Population Commission (NPC) and various development partners, including technical support from the Inner-City Fund (ICF) International. The NDHS provides essential health and demographic indicators that are up-to-date and nationally representative [23]. Ethics approval for the survey was granted by the Nigerian National Health Research Ethics Committee. Survey participants who were 18 years or older provided informed written consent, and consent was obtained from their parents or guardians if they were below 18. The 2018 NDHS, the sixth edition in its series, used a two-stage stratified cluster sampling method involving about 42,000 households and 1,400 clusters [23].

The survey conducted was successful in covering 40,427 households and 1,389 clusters. The selection criteria were predetermined, and validated questionnaires were employed to collect data from eligible men, women, and households [23]. The questionnaires were administered by interviewers, and 41,821 women between the ages of 15-49 participated, with 16,984 women from urban areas and 24,837 from rural areas. The eligible women showed a high response rate of 99.3%, with 99.2% from urban areas and 99.4% from rural areas. This study involved the analysis of data from a weighted sample of 21,553 mothers (8,440 in urban and 13,113 in rural areas) who provided complete information on ANC contacts for their most recent live childbirths that occurred within the five years preceding the survey [23]. We used the Children Recode (KR) dataset of the 2018 NDHS, which was completely anonymised before we accessed them, ensuring that there was no identifiable information about the survey participants. We received approved access to use the data, and no further ethical clearance was required to conduct this study. The 2018 NDHS sampling procedures, settings, questionnaires, and design have been previously released in a detailed report [23]. Access to the data used in this study is freely available online (https://dhsprogram.com/data/available-datasets.cfm) after obtaining the necessary approval from the Demographic and Health Survey (DHS) program.

### Variables

#### Outcome variable

This study’s primary outcome variable was ANC services utilisation, which we extracted from the 2018 NDHS data [23]. We defined ANC utilisation as having at least eight contacts, aligning with the updated WHO guideline and in line with practice in recent studies [1, 2, 14, 18–20, 22]. In this context, ANC services encompass pregnancy-related care administered to women and adolescent girls by qualified healthcare professionals [1, 2]. The ANC variable extracted from the 2018 NDHS was dichotomised into two categories: less than eight times contacts (< 8, suggesting underutilisation, coded as “0”) and at least eight times ANC contacts (≥ 8, indicating utilisation, coded as “1”). Mothers who responded with ‘don’t know’ or had missing information regarding the question “How many times did you receive antenatal care during this pregnancy?” were excluded from our analyses. As secondary outcomes, we equally investigated essential components recommended by the WHO for all expectant mothers attending ANC clinics and how their provision differs across rural and urban residences in Nigeria. These ANC components include treating intestinal parasites, administering iron tablets or syrup, administering tetanus toxoid vaccination, blood sample collection, monitoring blood pressure, and collecting urine samples [1, 2].

#### Explanatory variables

We adopted Andersen’s behavioural model [24] as the conceptual framework for selecting explanatory variables in this study, which is consistent with practice in previous studies [6, 13, 14]. Briefly, Andersen’s behavioural model is widely used as a framework in healthcare research, especially in studies examining factors influencing health service utilisation. In the context of ANC utilisation, the model considers three key components: predisposing factors (e.g., demographics), enabling factors (e.g., access to resources), and need factors (e.g., perceived health needs) [24]. Considering variables captured in the 2018 NDHS [23], we apply this model to assess how a range of factors are associated with a woman’s decision to seek ANC services. The model, as used in our study, is depicted in Figure 1. Following the example of previous studies [6, 13], we selected a total of twenty-two explanatory variables for inclusion in our analysis. We broadly grouped these variables into four categories: external environmental, predisposing, enabling, and need factors (Figure 1).

**Figure 1:**
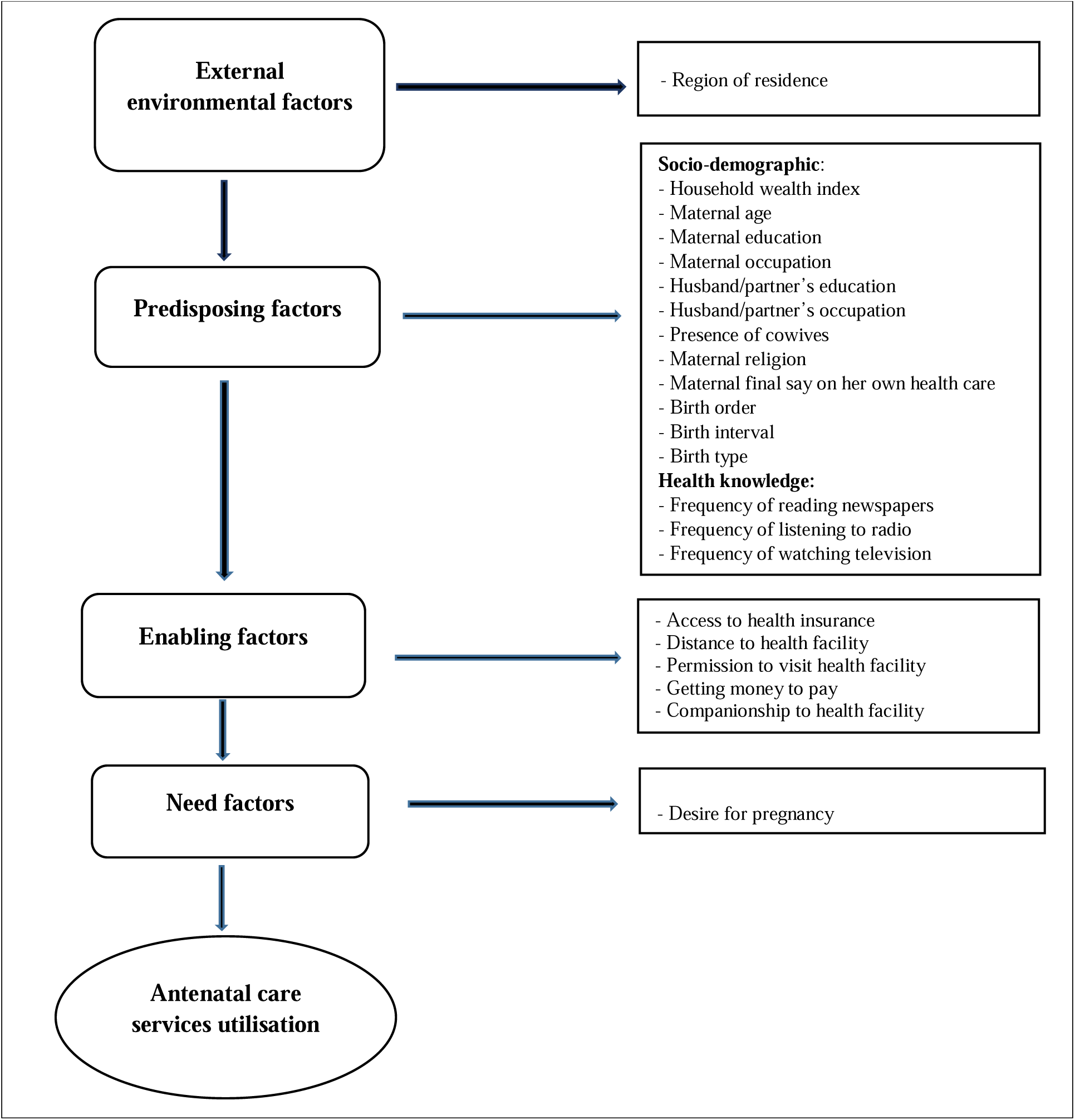
Theoretical framework to study factors antenatal care utilisation in Nigeria

External environmental factors include whether the study participants lived in an urban or rural area and which of Nigeria’s six geopolitical zones they reside in: ‘ North-West, North-East, North-Central, South-South, South-East, and South-West’ regions. We further classified the predisposing variables into health knowledge and socio-demographic factors. Health knowledge factors include media exposure variables such as frequency of reading newspapers/magazines, internet use, watching television, and listening to the radio. Socio-demographic factors, on the other hand, include the household wealth index, maternal religion, maternal age, maternal and husband/partner’s working status, and maternal and husband/partner’s education level. Additional socio-demographic factors considered were the mother’s final say on her health, ethnicity, preceding birth interval, birth order and birth type. Enabling factors encompass variables that can facilitate healthcare service utilisation, such as access to health insurance, companionship to a health facility, obtaining money to pay for health services, distance to a health facility, and getting permission to visit health facilities. Lastly, the ‘desire for pregnancy’ was assessed as a need factor. Table 1 provides additional information regarding specific categorisations of these dependent variables.

**Table 1:**
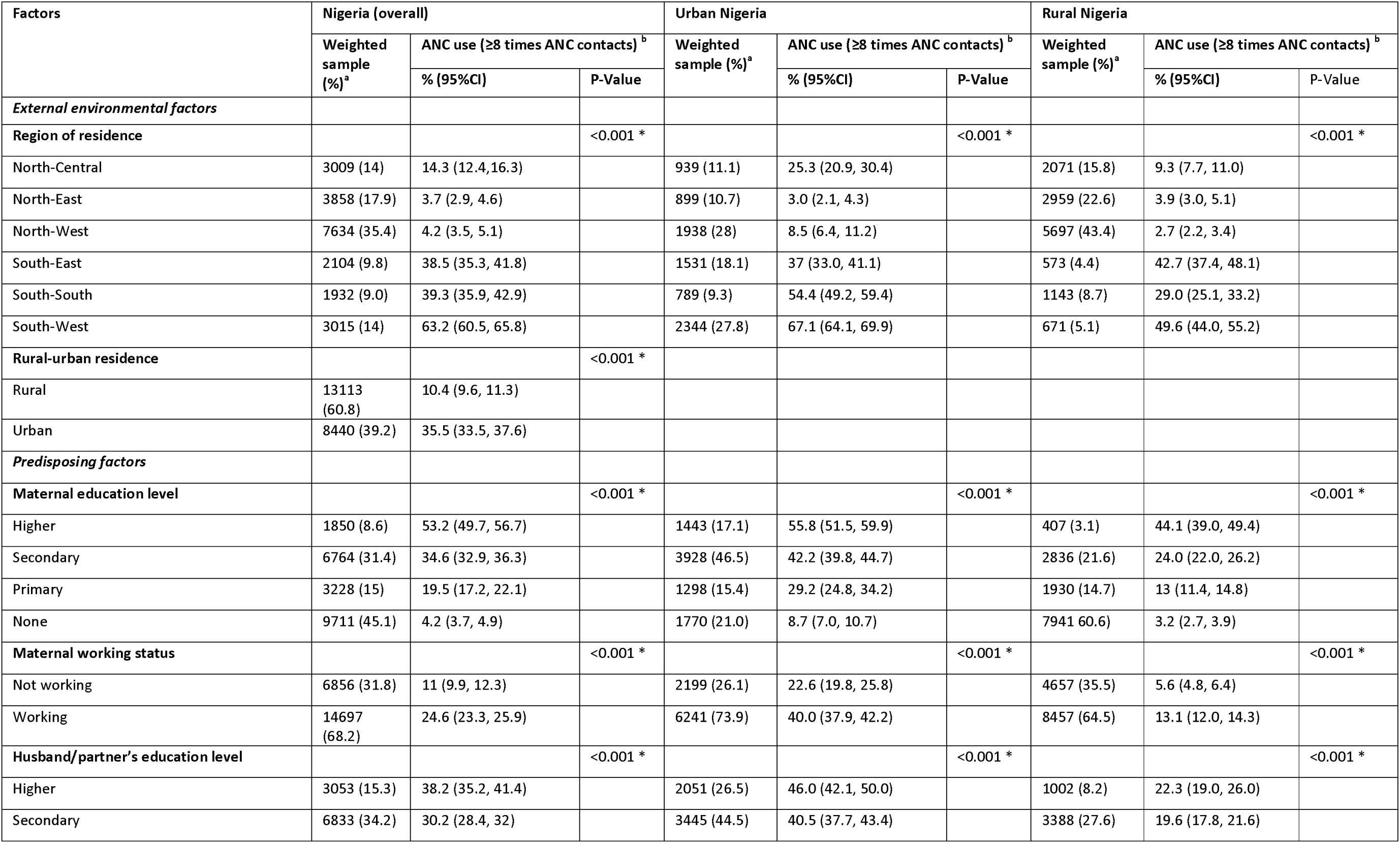
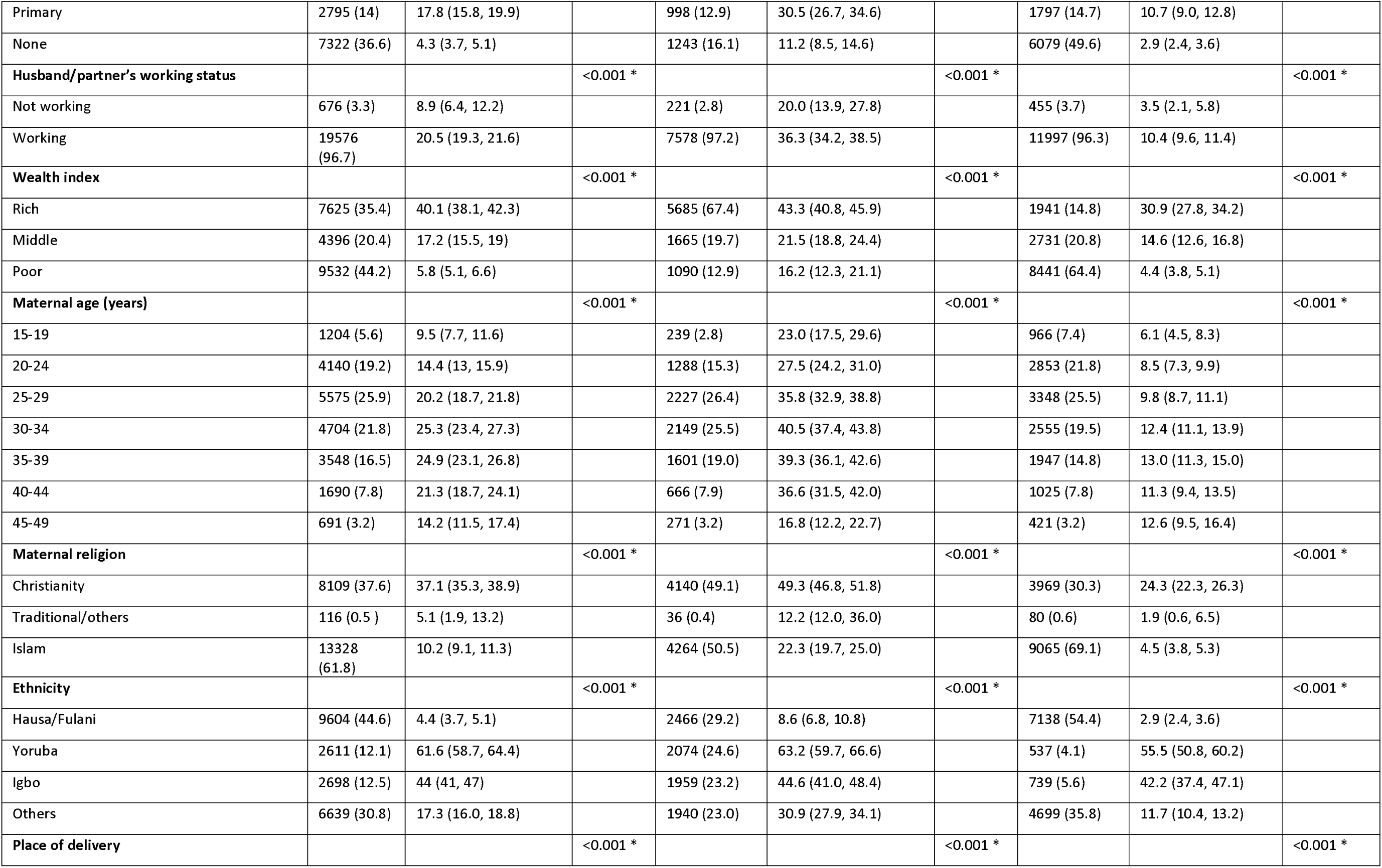
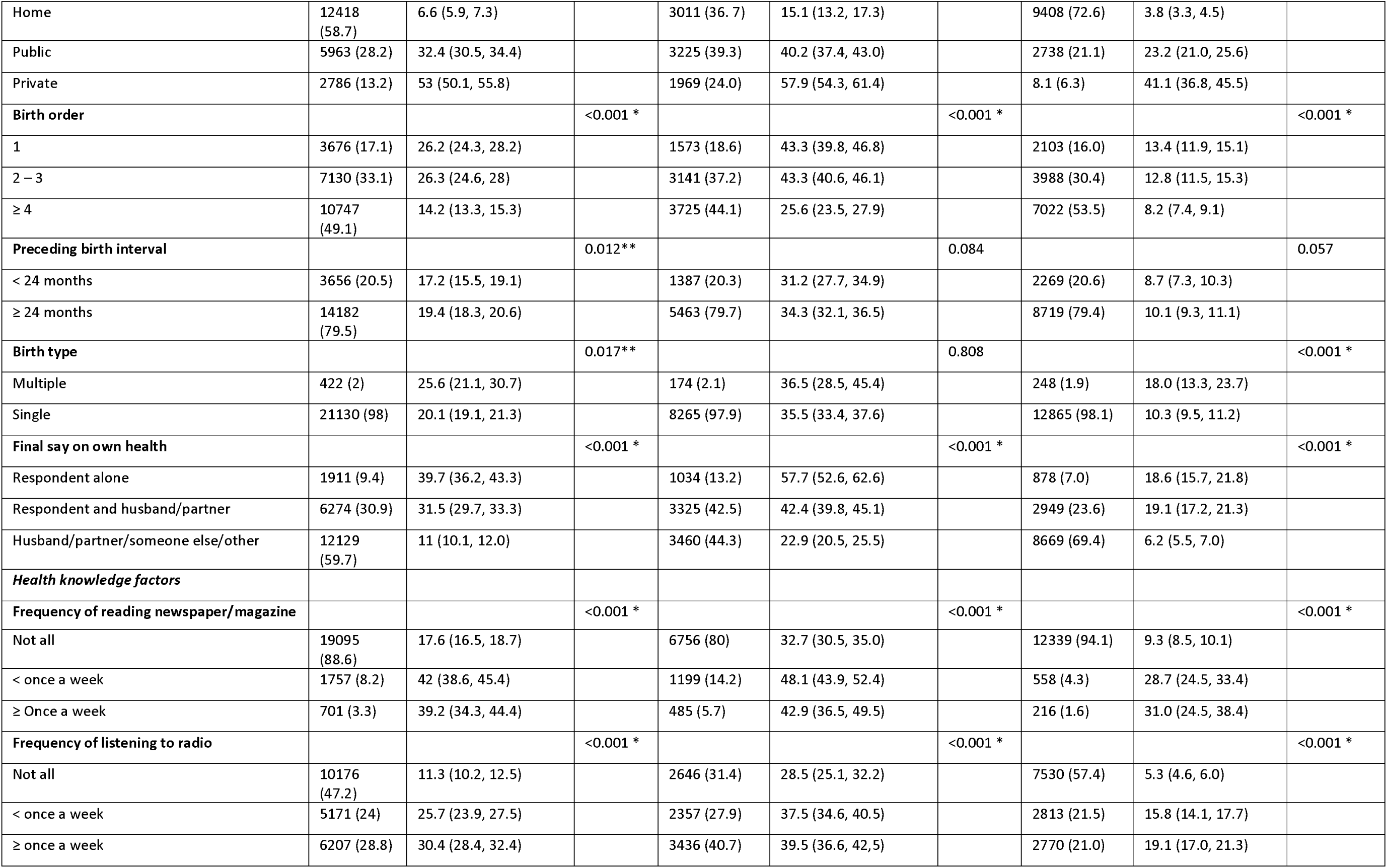
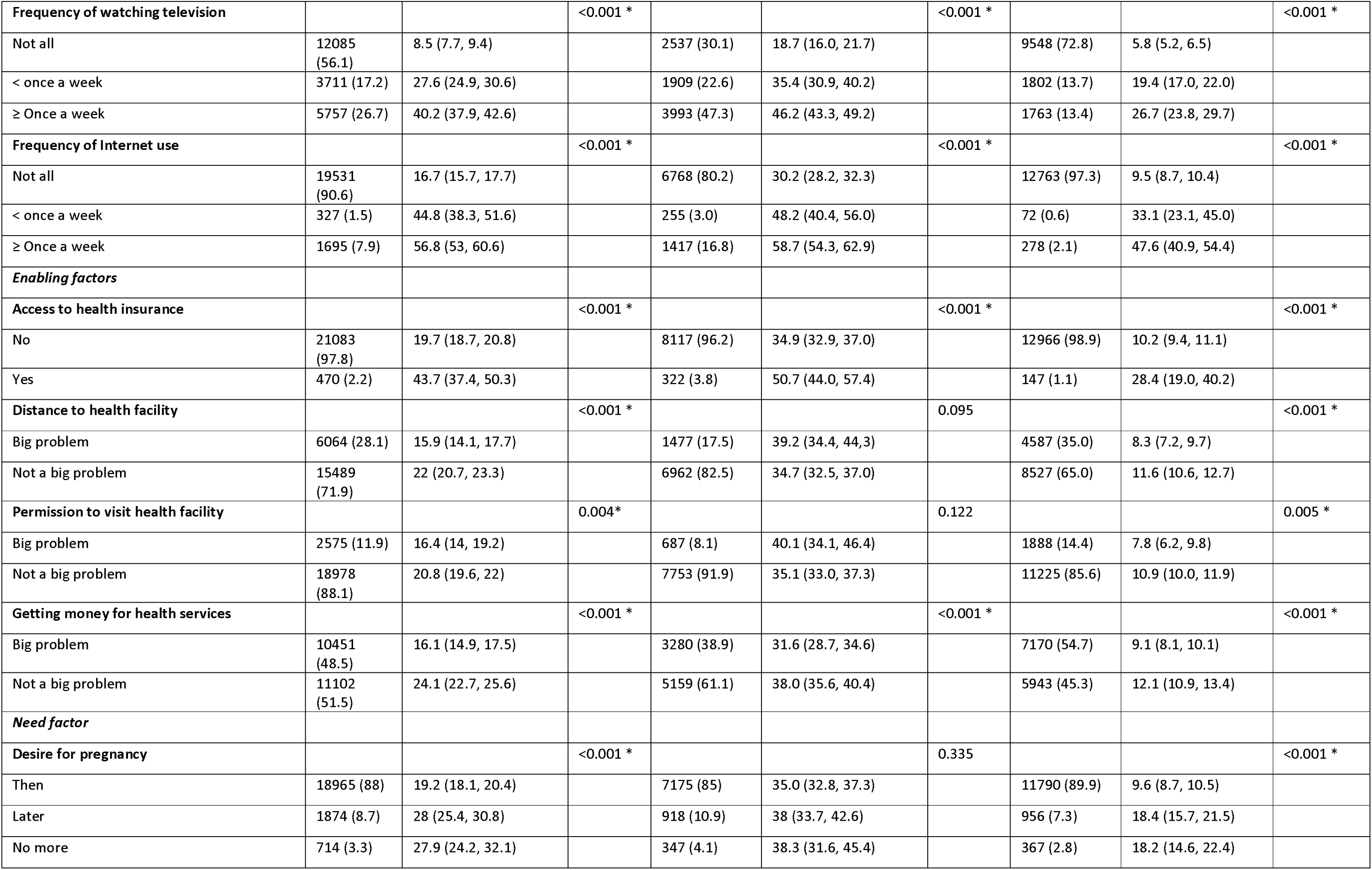
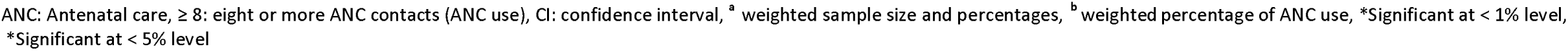
Sample characteristics and antenatal care utilisation of ≥ 8 contacts in Nigeria by rural and urban residences.

| Factors | Nigeria (overall) |  |  | Urban Nigeria |  |  | Rural Nigeria |  |  |
| --- | --- | --- | --- | --- | --- | --- | --- | --- | --- |
|  | Weighted sample (%)<sup>a</sup> | ANC use (≥8 times ANC contacts)<sup>b</sup> |  | Weighted sample (%)<sup>a</sup> | ANC use (≥8 times ANC contacts)<sup>b</sup> |  | Weighted sample (%)<sup>a</sup> | ANC use (≥8 times ANC contacts)<sup>b</sup> |  |
|  |  | % (95%CI) | P-Value |  | % (95%CI) | P-Value |  | % (95%CI) | P-Value |
| *External environmental factors* |  |  |  |  |  |  |  |  |  |
| **Region of residence** |  |  | <0.001 * |  |  | <0.001 * |  |  | <0.001 * |
| North-Central | 3009 (14) | 14.3 (12.4, 16.3) |  | 939 (11.1) | 25.3 (20.9, 30.4) |  | 2071 (15.8) | 9.3 (7.7, 11.0) |  |
| North-East | 3858 (17.9) | 3.7 (2.9, 4.6) |  | 899 (10.7) | 3.0 (2.1, 4.3) |  | 2959 (22.6) | 3.9 (3.0, 5.1) |  |
| North-West | 7634 (35.4) | 4.2 (3.5, 5.1) |  | 1938 (28) | 8.5 (6.4, 11.2) |  | 5697 (43.4) | 2.7 (2.2, 3.4) |  |
| South-East | 2104 (9.8) | 38.5 (35.3, 41.8) |  | 1531 (18.1) | 37 (33.0, 41.1) |  | 573 (4.4) | 42.7 (37.4, 48.1) |  |
| South-South | 1932 (9.0) | 39.3 (35.9, 42.9) |  | 789 (9.3) | 54.4 (49.2, 59.4) |  | 1143 (8.7) | 29.0 (25.1, 33.2) |  |
| South-West | 3015 (14) | 63.2 (60.5, 65.8) |  | 2344 (27.8) | 67.1 (64.1, 69.9) |  | 671 (5.1) | 49.6 (44.0, 55.2) |  |
| **Rural-urban residence** |  |  | <0.001 * |  |  |  |  |  |  |
| Rural | 13113 (60.8) | 10.4 (9.6, 11.3) |  |  |  |  |  |  |  |
| Urban | 8440 (39.2) | 35.5 (33.5, 37.6) |  |  |  |  |  |  |  |
| *Predisposing factors* |  |  |  |  |  |  |  |  |  |
| **Maternal education level** |  |  | <0.001 * |  |  | <0.001 * |  |  | <0.001 * |
| Higher | 1850 (8.6) | 53.2 (49.7, 56.7) |  | 1443 (17.1) | 55.8 (51.5, 59.9) |  | 407 (3.1) | 44.1 (39.0, 49.4) |  |
| Secondary | 6764 (31.4) | 34.6 (32.9, 36.3) |  | 3928 (46.5) | 42.2 (39.8, 44.7) |  | 2836 (21.6) | 24.0 (22.0, 26.2) |  |
| Primary | 3228 (15) | 19.5 (17.2, 22.1) |  | 1298 (15.4) | 29.2 (24.8, 34.2) |  | 1930 (14.7) | 13 (11.4, 14.8) |  |
| None | 9711 (45.1) | 4.2 (3.7, 4.9) |  | 1770 (21.0) | 8.7 (7.0, 10.7) |  | 7941 (60.6) | 3.2 (2.7, 3.9) |  |
| **Maternal working status** |  |  | <0.001 * |  |  | <0.001 * |  |  | <0.001 * |
| Not working | 6856 (31.8) | 11 (9.9, 12.3) |  | 2199 (26.1) | 22.6 (19.8, 25.8) |  | 4657 (35.5) | 5.6 (4.8, 6.4) |  |
| Working | 14697 (68.2) | 24.6 (23.3, 25.9) |  | 6241 (73.9) | 40.0 (37.9, 42.2) |  | 8457 (64.5) | 13.1 (12.0, 14.3) |  |
| **Husband/partner's education level** |  |  | <0.001 * |  |  | <0.001 * |  |  | <0.001 * |
| Higher | 3053 (15.3) | 38.2 (35.2, 41.4) |  | 2051 (26.5) | 46.0 (42.1, 50.0) |  | 1002 (8.2) | 22.3 (19.0, 26.0) |  |
| Secondary | 6833 (34.2) | 30.2 (28.4, 32) |  | 3445 (44.5) | 40.5 (37.7, 43.4) |  | 3388 (27.6) | 19.6 (17.8, 21.6) |  |
| Primary | 2795 (14) | 17.8 (15.8, 19.9) |  | 998 (12.9) | 30.5 (26.7, 34.6) |  | 1797 (14.7) | 10.7 (9.0, 12.8) |  |
| None | 7322 (36.6) | 4.3 (3.7, 5.1) |  | 1243 (16.1) | 11.2 (8.5, 14.6) |  | 6079 (49.6) | 2.9 (2.4, 3.6) |  |
| **Husband/partner's working status** |  |  | <0.001 * |  |  | <0.001 * |  |  | <0.001 * |
| Not working | 676 (3.3) | 8.9 (6.4, 12.2) |  | 221 (2.8) | 20.0 (13.9, 27.8) |  | 455 (3.7) | 3.5 (2.1, 5.8) |  |
| Working | 19576 (96.7) | 20.5 (19.3, 21.6) |  | 7578 (97.2) | 36.3 (34.2, 38.5) |  | 11997 (96.3) | 10.4 (9.6, 11.4) |  |
| **Wealth index** |  |  | <0.001 * |  |  | <0.001 * |  |  | <0.001 * |
| Rich | 7625 (35.4) | 40.1 (38.1, 42.3) |  | 5685 (67.4) | 43.3 (40.8, 45.9) |  | 1941 (14.8) | 30.9 (27.8, 34.2) |  |
| Middle | 4396 (20.4) | 17.2 (15.5, 19) |  | 1665 (19.7) | 21.5 (18.8, 24.4) |  | 2731 (20.8) | 14.6 (12.6, 16.8) |  |
| Poor | 9532 (44.2) | 5.8 (5.1, 6.6) |  | 1090 (12.9) | 16.2 (12.3, 21.1) |  | 8441 (64.4) | 4.4 (3.8, 5.1) |  |
| **Maternal age (years)** |  |  | <0.001 * |  |  | <0.001 * |  |  | <0.001 * |
| 15-19 | 1204 (5.6) | 9.5 (7.7, 11.6) |  | 239 (2.8) | 23.0 (17.5, 29.6) |  | 966 (7.4) | 6.1 (4.5, 8.3) |  |
| 20-24 | 4140 (19.2) | 14.4 (13, 15.9) |  | 1288 (15.3) | 27.5 (24.2, 31.0) |  | 2853 (21.8) | 8.5 (7.3, 9.9) |  |
| 25-29 | 5575 (25.9) | 20.2 (18.7, 21.8) |  | 2227 (26.4) | 35.8 (32.9, 38.8) |  | 3348 (25.5) | 9.8 (8.7, 11.1) |  |
| 30-34 | 4704 (21.8) | 25.3 (23.4, 27.3) |  | 2149 (25.5) | 40.5 (37.4, 43.8) |  | 2555 (19.5) | 12.4 (11.1, 13.9) |  |
| 35-39 | 3548 (16.5) | 24.9 (23.1, 26.8) |  | 1601 (19.0) | 39.3 (36.1, 42.6) |  | 1947 (14.8) | 13.0 (11.3, 15.0) |  |
| 40-44 | 1690 (7.8) | 21.3 (18.7, 24.1) |  | 666 (7.9) | 36.6 (31.5, 42.0) |  | 1025 (7.8) | 11.3 (9.4, 13.5) |  |
| 45-49 | 691 (3.2) | 14.2 (11.5, 17.4) |  | 271 (3.2) | 16.8 (12.2, 22.7) |  | 421 (3.2) | 12.6 (9.5, 16.4) |  |
| **Maternal religion** |  |  | <0.001 * |  |  | <0.001 * |  |  | <0.001 * |
| Christianity | 8109 (37.6) | 37.1 (35.3, 38.9) |  | 4140 (49.1) | 49.3 (46.8, 51.8) |  | 3969 (30.3) | 24.3 (22.3, 26.3) |  |
| Traditional/others | 116 (0.5 ) | 5.1 (1.9, 13.2) |  | 36 (0.4) | 12.2 (12.0, 36.0) |  | 80 (0.6) | 1.9 (0.6, 6.5) |  |
| Islam | 13328 (61.8) | 10.2 (9.1, 11.3) |  | 4264 (50.5) | 22.3 (19.7, 25.0) |  | 9065 (69.1) | 4.5 (3.8, 5.3) |  |
| **Ethnicity** |  |  | <0.001 * |  |  | <0.001 * |  |  | <0.001 * |
| Hausa/Fulani | 9604 (44.6) | 4.4 (3.7, 5.1) |  | 2466 (29.2) | 8.6 (6.8, 10.8) |  | 7138 (54.4) | 2.9 (2.4, 3.6) |  |
| Yoruba | 2611 (12.1) | 61.6 (58.7, 64.4) |  | 2074 (24.6) | 63.2 (59.7, 66.6) |  | 537 (4.1) | 55.5 (50.8, 60.2) |  |
| Igbo | 2698 (12.5) | 44 (41, 47) |  | 1959 (23.2) | 44.6 (41.0, 48.4) |  | 739 (5.6) | 42.2 (37.4, 47.1) |  |
| Others | 6639 (30.8) | 17.3 (16.0, 18.8) |  | 1940 (23.0) | 30.9 (27.9, 34.1) |  | 4699 (35.8) | 11.7 (10.4, 13.2) |  |
| **Place of delivery** |  |  | <0.001 * |  |  | <0.001 * |  |  | <0.001 * |
| Home | 12418<br>(58.7) | 6.6 (5.9, 7.3) |  | 3011 (36.7) | 15.1 (13.2, 17.3) |  | 9408 (72.6) | 3.8 (3.3, 4.5) |  |
| Public | 5963 (28.2) | 32.4 (30.5, 34.4) |  | 3225 (39.3) | 40.2 (37.4, 43.0) |  | 2738 (21.1) | 23.2 (21.0, 25.6) |  |
| Private | 2786 (13.2) | 53 (50.1, 55.8) |  | 1969 (24.0) | 57.9 (54.3, 61.4) |  | 8.1 (6.3) | 41.1 (36.8, 45.5) |  |
| **Birth order** |  |  | <0.001 * |  |  | <0.001 * |  |  | <0.001 * |
| 1 | 3676 (17.1) | 26.2 (24.3, 28.2) |  | 1573 (18.6) | 43.3 (39.8, 46.8) |  | 2103 (16.0) | 13.4 (11.9, 15.1) |  |
| 2 – 3 | 7130 (33.1) | 26.3 (24.6, 28) |  | 3141 (37.2) | 43.3 (40.6, 46.1) |  | 3988 (30.4) | 12.8 (11.5, 15.3) |  |
| ≥ 4 | 10747<br>(49.1) | 14.2 (13.3, 15.3) |  | 3725 (44.1) | 25.6 (23.5, 27.9) |  | 7022 (53.5) | 8.2 (7.4, 9.1) |  |
| **Preceding birth interval** |  |  | 0.012** |  |  | 0.084 |  |  | 0.057 |
| < 24 months | 3656 (20.5) | 17.2 (15.5, 19.1) |  | 1387 (20.3) | 31.2 (27.7, 34.9) |  | 2269 (20.6) | 8.7 (7.3, 10.3) |  |
| ≥ 24 months | 14182<br>(79.5) | 19.4 (18.3, 20.6) |  | 5463 (79.7) | 34.3 (32.1, 36.5) |  | 8719 (79.4) | 10.1 (9.3, 11.1) |  |
| **Birth type** |  |  | 0.017** |  |  | 0.808 |  |  | <0.001 * |
| Multiple | 422 (2) | 25.6 (21.1, 30.7) |  | 174 (2.1) | 36.5 (28.5, 45.4) |  | 248 (1.9) | 18.0 (13.3, 23.7) |  |
| Single | 21130 (98) | 20.1 (19.1, 21.3) |  | 8265 (97.9) | 35.5 (33.4, 37.6) |  | 12865 (98.1) | 10.3 (9.5, 11.2) |  |
| **Final say on own health** |  |  | <0.001 * |  |  | <0.001 * |  |  | <0.001 * |
| Respondent alone | 1911 (9.4) | 39.7 (36.2, 43.3) |  | 1034 (13.2) | 57.7 (52.6, 62.6) |  | 878 (7.0) | 18.6 (15.7, 21.8) |  |
| Respondent and husband/partner | 6274 (30.9) | 31.5 (29.7, 33.3) |  | 3325 (42.5) | 42.4 (39.8, 45.1) |  | 2949 (23.6) | 19.1 (17.2, 21.3) |  |
| Husband/partner/someone else/other | 12129<br>(59.7) | 11 (10.1, 12.0) |  | 3460 (44.3) | 22.9 (20.5, 25.5) |  | 8669 (69.4) | 6.2 (5.5, 7.0) |  |
| **Health knowledge factors** |  |  |  |  |  |  |  |  |  |
| **Frequency of reading newspaper/magazine** |  |  | <0.001 * |  |  | <0.001 * |  |  | <0.001 * |
| Not all | 19095<br>(88.6) | 17.6 (16.5, 18.7) |  | 6756 (80) | 32.7 (30.5, 35.0) |  | 12339 (94.1) | 9.3 (8.5, 10.1) |  |
| < once a week | 1757 (8.2) | 42 (38.6, 45.4) |  | 1199 (14.2) | 48.1 (43.9, 52.4) |  | 558 (4.3) | 28.7 (24.5, 33.4) |  |
| ≥ Once a week | 701 (3.3) | 39.2 (34.3, 44.4) |  | 485 (5.7) | 42.9 (36.5, 49.5) |  | 216 (1.6) | 31.0 (24.5, 38.4) |  |
| **Frequency of listening to radio** |  |  | <0.001 * |  |  | <0.001 * |  |  | <0.001 * |
| Not all | 10176<br>(47.2) | 11.3 (10.2, 12.5) |  | 2646 (31.4) | 28.5 (25.1, 32.2) |  | 7530 (57.4) | 5.3 (4.6, 6.0) |  |
| < once a week | 5171 (24) | 25.7 (23.9, 27.5) |  | 2357 (27.9) | 37.5 (34.6, 40.5) |  | 2813 (21.5) | 15.8 (14.1, 17.7) |  |
| ≥ once a week | 6207 (28.8) | 30.4 (28.4, 32.4) |  | 3436 (40.7) | 39.5 (36.6, 42.5) |  | 2770 (21.0) | 19.1 (17.0, 21.3) |  |
| **Frequency of watching television** |  |  | <0.001 * |  |  | <0.001 * |  |  | <0.001 * |
| Not all | 12085<br>(56.1) | 8.5 (7.7, 9.4) |  | 2537 (30.1) | 18.7 (16.0, 21.7) |  | 9548 (72.8) | 5.8 (5.2, 6.5) |  |
| < once a week | 3711 (17.2) | 27.6 (24.9, 30.6) |  | 1909 (22.6) | 35.4 (30.9, 40.2) |  | 1802 (13.7) | 19.4 (17.0, 22.0) |  |
| ≥ Once a week | 5757 (26.7) | 40.2 (37.9, 42.6) |  | 3993 (47.3) | 46.2 (43.3, 49.2) |  | 1763 (13.4) | 26.7 (23.8, 29.7) |  |
| **Frequency of Internet use** |  |  | <0.001 * |  |  | <0.001 * |  |  | <0.001 * |
| Not all | 19531<br>(90.6) | 16.7 (15.7, 17.7) |  | 6768 (80.2) | 30.2 (28.2, 32.3) |  | 12763 (97.3) | 9.5 (8.7, 10.4) |  |
| < once a week | 327 (1.5) | 44.8 (38.3, 51.6) |  | 255 (3.0) | 48.2 (40.4, 56.0) |  | 72 (0.6) | 33.1 (23.1, 45.0) |  |
| ≥ Once a week | 1695 (7.9) | 56.8 (53, 60.6) |  | 1417 (16.8) | 58.7 (54.3, 62.9) |  | 278 (2.1) | 47.6 (40.9, 54.4) |  |
| **Enabling factors** |  |  |  |  |  |  |  |  |  |
| **Access to health insurance** |  |  | <0.001 * |  |  | <0.001 * |  |  | <0.001 * |
| No | 21083<br>(97.8) | 19.7 (18.7, 20.8) |  | 8117 (96.2) | 34.9 (32.9, 37.0) |  | 12966 (98.9) | 10.2 (9.4, 11.1) |  |
| Yes | 470 (2.2) | 43.7 (37.4, 50.3) |  | 322 (3.8) | 50.7 (44.0, 57.4) |  | 147 (1.1) | 28.4 (19.0, 40.2) |  |
| **Distance to health facility** |  |  | <0.001 * |  |  | 0.095 |  |  | <0.001 * |
| Big problem | 6064 (28.1) | 15.9 (14.1, 17.7) |  | 1477 (17.5) | 39.2 (34.4, 44.3) |  | 4587 (35.0) | 8.3 (7.2, 9.7) |  |
| Not a big problem | 15489<br>(71.9) | 22 (20.7, 23.3) |  | 6962 (82.5) | 34.7 (32.5, 37.0) |  | 8527 (65.0) | 11.6 (10.6, 12.7) |  |
| **Permission to visit health facility** |  |  | 0.004 * |  |  | 0.122 |  |  | 0.005 * |
| Big problem | 2575 (11.9) | 16.4 (14, 19.2) |  | 687 (8.1) | 40.1 (34.1, 46.4) |  | 1888 (14.4) | 7.8 (6.2, 9.8) |  |
| Not a big problem | 18978<br>(88.1) | 20.8 (19.6, 22) |  | 7753 (91.9) | 35.1 (33.0, 37.3) |  | 11225 (85.6) | 10.9 (10.0, 11.9) |  |
| **Getting money for health services** |  |  | <0.001 * |  |  | <0.001 * |  |  | <0.001 * |
| Big problem | 10451<br>(48.5) | 16.1 (14.9, 17.5) |  | 3280 (38.9) | 31.6 (28.7, 34.6) |  | 7170 (54.7) | 9.1 (8.1, 10.1) |  |
| Not a big problem | 11102<br>(51.5) | 24.1 (22.7, 25.6) |  | 5159 (61.1) | 38.0 (35.6, 40.4) |  | 5943 (45.3) | 12.1 (10.9, 13.4) |  |
| **Need factor** |  |  |  |  |  |  |  |  |  |
| **Desire for pregnancy** |  |  | <0.001 * |  |  | 0.335 |  |  | <0.001 * |
| Then | 18965 (88) | 19.2 (18.1, 20.4) |  | 7175 (85) | 35.0 (32.8, 37.3) |  | 11790 (89.9) | 9.6 (8.7, 10.5) |  |
| Later | 1874 (8.7) | 28 (25.4, 30.8) |  | 918 (10.9) | 38 (33.7, 42.6) |  | 956 (7.3) | 18.4 (15.7, 21.5) |  |
| No more | 714 (3.3) | 27.9 (24.2, 32.1) |  | 347 (4.1) | 38.3 (31.6, 45.4) |  | 367 (2.8) | 18.2 (14.6, 22.4) |  |

ANC: Antenatal care, $\geq 8$ : eight or more ANC contacts (ANC use), CI: confidence interval, <sup>a</sup> weighted sample size and percentages, <sup>b</sup> weighted percentage of ANC use, \*Significant at < 1% level, \*Significant at < 5% level

#### Statistical analysis

We assess the unadjusted and adjusted association between ANC utilisation (proportion of mothers with ≥ 8 ANC contacts) with explanatory variables of interest, first for the overall Nigerian population and then separately for rural and urban residences in the country. We estimated ANC use and its 95% confidence interval (CI) through frequency tabulation and assessed the unadjusted relationship between ANC use and the explanatory variables by using a Chi-Square test. We conducted multivariable binary logistic regression analyses to identify factors significantly associated with ANC utilisation in Nigeria following adjustment for confounders or other predictor variables. In our model-building process, we only included variables that showed statistical significance (p < 0.05 in the Chi-Square test) in the initial model of the multivariable regression analyses. The backward elimination procedure helped to identify significant variables associated with ANC use at a 5% significance level (p < 0.05). We reported the adjusted odds ratios (AOR), their 95% confidence intervals (CI) and p-values for the variables retained in the final model. Moreover, we considered potential confounders or factors previously reported in studies to ensure we didn’t miss any significant variables. We also estimated the receipt of essential ANC components, first for the overall Nigerian population and thereafter, separately for rural and urban residences. We used the Statistical Package for Social Sciences (SPSS), version 21 (IBM SPSS Statistics for Windows, Version 21.0. Armonk, NY: IBM Corp) for data management and analyses. In all analyses, we used the Complex Sample analysis method, considering the stratified design and the sampling weight of the 2018 NDHS.

#### Patient and public involvement statement

This research study utilised pre-existing, fully anonymised data. As a secondary data analysis, patients were not involved in the study. The survey design and execution, however, involved gathering data from respondents and the participation of relevant stakeholders, such as government and non-government organisations, in the survey implementation [23].

## Results

### Sample characteristics for the overall, rural, and urban residences

Table 1 details the characteristics of the sample examined in our study, including data from rural and urban residences in Nigeria. Our study included a total weighted sample of 21,553 mothers aged 15 – 49 with information about their ANC contacts. Of these, 60.8% (13113) were from rural residences, and 39.2% (8440) were from urban areas. In Nigeria, 5.6% of participants were teenagers. The proportion of teenage mothers was lower in urban areas (2.8%) compared to rural areas (7.4%). A high proportion of participants (45.1%) had no formal education, with this trend being more pronounced in rural areas (60.6%) than in urban areas (21.0%). Similarly, most participants were from poor households (44.2%), with rural areas having a substantially higher percentage (64.4%) compared to urban areas (12.9%). Approximately 60% of mothers delivered their babies at home in Nigeria—36.7% in urban residences and over 70% in rural areas. Internet use at least once a week was low at 7.9%, with rural areas having considerably low usage (2.1%) compared to urban areas (16.8%). Access to health insurance coverage was also low at 2.2% in Nigeria—3.8% in urban and 1.1% in rural areas. Nearly half of women (48.5%) reported having a big challenge with getting money for health services. This challenge was more prominent in rural residences (54.7%) than urban residences (38.9%).

### Eight or more ANC utilisation in Nigeria by rural and urban residences

A total of 4,366 mothers (20.3%, 95% CI: 19.2, 21.4%, P1<10.001) reported having at least eight (≥ 8) ANC contacts in Nigeria (Table 1 and Figure 2). Urban mothers notably recorded higher ANC utilisation (35.5%, 95% CI: 33.5, 37.6%, P1<10.001) than their rural counterparts (10.4%, 95% CI: 9.6, 11.3%, P1<10.001, Figure 2). At the regional level, the South-West geopolitical zone had the highest ANC utilisation at 63.2% (95% CI: 60.5, 65.8%, P1<10.001) in Nigeria, consistent across rural (49.6%, 95% CI: 44.0, 55.2%, P1<10.001) and urban (67.1%, 95% CI: 64.1, 69.9%, P1<10.001) residences (Figure 3). In contrast, the North-East region recorded the lowest ANC use in Nigeria (3.7%, 95% CI: 2.9, 4.6%, P1<10.001), and urban Nigeria (3.0%, 95% CI: 2.1, 4.3%, P1<10.001), while the North-West region had the lowest in rural Nigeria (2.7%, 95% CI: 2.2, 3.4%, P1<10.001) as summarised in Figure 3.

**Figure 2:**
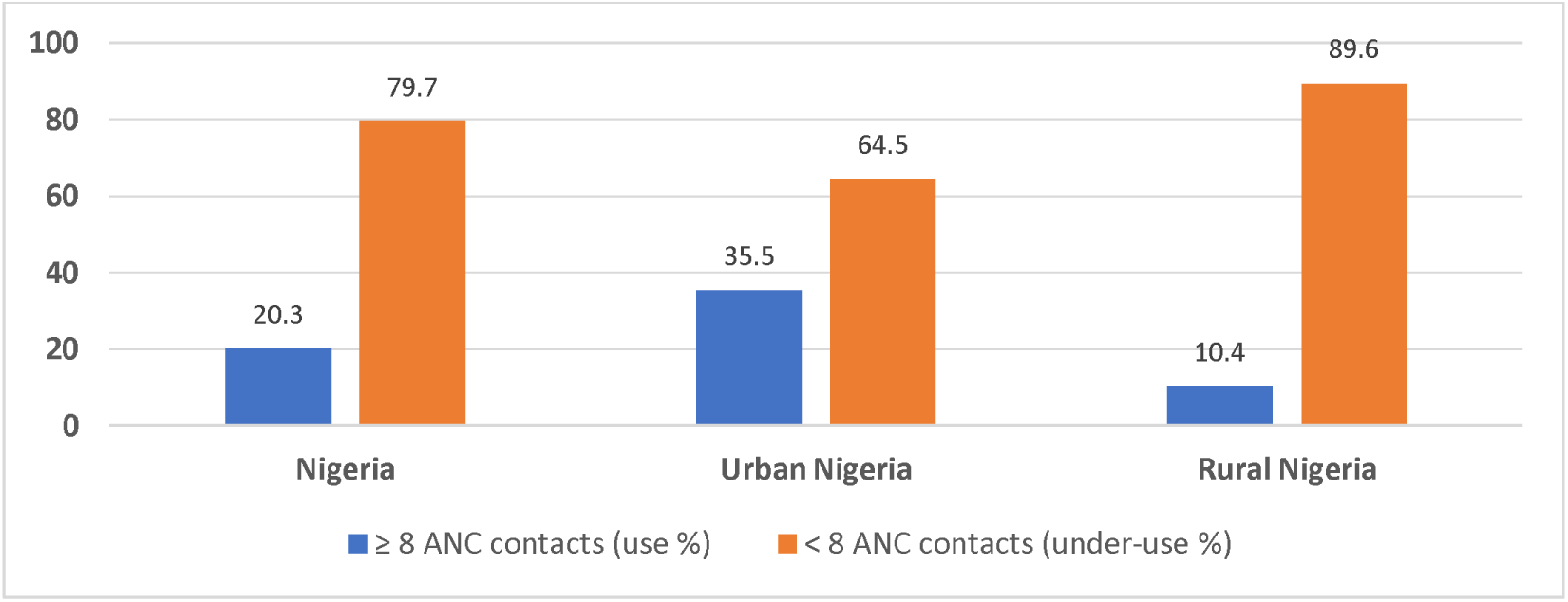
ANC use and under-use in Nigeria by rural-urban residences

**Figure 3:**
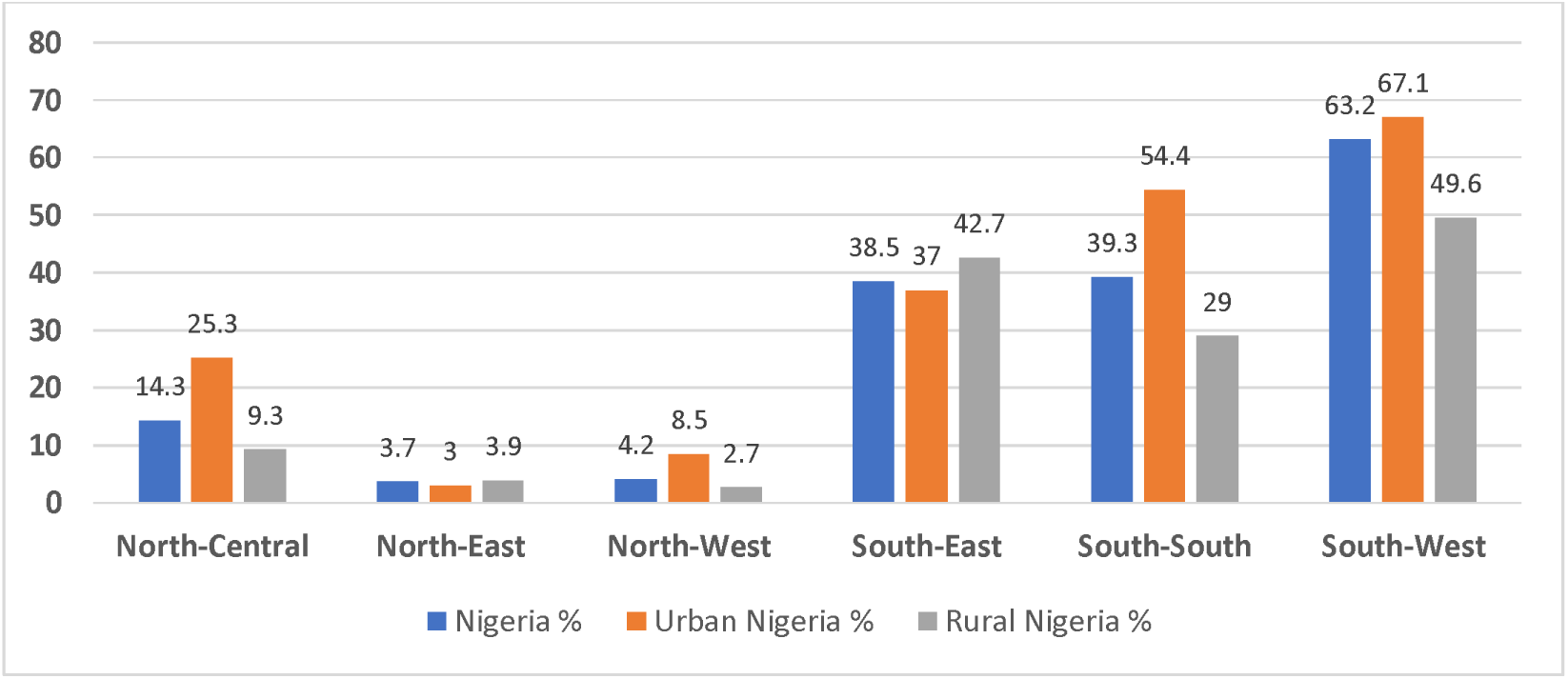
ANC use in Nigeria’s geopolitical zones by rural-urban residences

Considering predisposing factors, all socio-demographic and health knowledge variables were significantly associated with ANC utilisation in Nigeria, rural or urban residences notwithstanding (Table 1). However, there were two exceptions, namely ‘preceding birth interval’ (significantly associated with ANC use only in the overall population) and ‘birth type’, which did not reach statistical significance in urban areas (Table 1). Among the enabling factors, distance to and permission to visit health facilities were not statistically significant in urban areas. The ‘need factor’ also showed a similar relationship with ANC utilisation, where the ‘desire for pregnancy’ was significant in the overall population and rural residences but not in urban areas. Table 1 provides additional details on ANC use for all the predictor variables across Nigeria’s overall, rural, and urban residences.

### ANC utilisation based on the focused model (four or more ANC contacts)

These results enable a comparison with the 2013 study[13]. Based on the 2018 NDHS, a total of 12456 (57.8%) mothers had four or more (≥4) ANC contacts in Nigeria—6418 (76.1%) in urban areas and 6038 (46.0%) in rural areas. Conversely, 9097 (42.2%) had less than four ANC contacts (underuse) in Nigeria—2021 (23.9%) in urban and 7075 (54%) in rural settings. Moreover, 5,336 mothers (24.4%) did not have ANC contact at all in the overall Nigerian population—878 (10.1%) were in urban residences, and 4,458 (33.8%) were in rural residences. Additionally, 574 (2.6%) mothers had only one ANC contact: 158 [1.8%] in urban and 416 [3.2%] in rural areas, 970 (4.4%) had two contacts: 264 [3.0%] in urban and 706 [5.4%] in rural areas, 2216 (10.1%) had three contacts: 722 [8.3%] in urban and 1494 [11.3%] in rural areas in Nigeria.

### Components of ANC received in Nigeria across rural-urban settings

Table 2 presents a comprehensive breakdown of ANC components in Nigeria, including the provision of iron tablets or syrup, tetanus injections, drugs for intestinal parasites, blood pressure, urine samples, and blood sample assessments during pregnancy, with a focus on the rural-urban divide. In the overall Nigerian context, 69.3% received iron tablets or syrup during pregnancy, 70.0% had tetanus injections, and 16.7% were administered drugs for intestinal parasites. Blood pressure, urine samples, and blood sample assessments were conducted for 93.9%, 86.4%, and 87.6% of pregnant women, respectively. Examining urban and rural disparities reveals that urban residents generally exhibit higher percentages across all components of ANC, with considerable differences observed in the provision of iron tablets or syrups (80.1% urban vs. 62.2% rural) and tetanus injections (84.3% urban vs. 60.6% rural). The gap is narrower for other components but consistently favours urban settings (Table 2).

**Table 2:** Components of ANC received in Nigeria disaggregated by rural and urban residences.

| S/N | Factors | Overall (Nigeria) |  |  | Urban Nigeria |  |  | Rural Nigeria |  |  |
| --- | --- | --- | --- | --- | --- | --- | --- | --- | --- | --- |
|  |  | N<sup>a</sup> | %<sup>b</sup> (95%CI) | P-value<sup>c</sup> | N<sup>a</sup> | %<sup>b</sup> (95%CI) | P-value<sup>c</sup> | N<sup>a</sup> | %<sup>b</sup> (95%CI) | P-value<sup>c</sup> |
| 1 | Given or bought iron tablets/syrup |  |  |  |  |  |  |  |  |  |
|  | Yes | 15189 | 69.3 (67.7, 70.9) | < 0.001* | 6978 | 80.1 (77.8, 82.2) | < 0.001* | 8211 | 62.2 (60.0, 64.4) | < 0.001* |
| 2 | Received tetanus injections before birth |  |  |  |  |  |  |  |  |  |
|  | Yes | 15267 | 70.0 (68.4, 71.5) | < 0.001* | 7290 | 84.3 (82.6, 85.9) | < 0.001* | 7977 | 60.6 (58.4, 62.8) | < 0.001* |
| 3 | Had drugs for intestinal parasites during pregnancy |  |  |  |  |  |  |  |  |  |
|  | Yes | 3666 | 16.7 (15.9, 17.6) | < 0.001* | 1624 | 18.6 (17.4, 19.9) | < 0.001* | 2042 | 15.5 (14.4, 16.7) | < 0.001* |
| 4 | During pregnancy: blood pressure taken |  |  |  |  |  |  |  |  |  |
|  | Yes | 15566 | 93.9 (93.2, 94.5) | < 0.001* | 7525 | 96.1 (95.1, 96.9) | < 0.001* | 8042 | 92.0 (91.0, 92.9) | < 0.001* |
| 5 | During pregnancy: urine sample taken |  |  |  |  |  |  |  |  |  |
|  | Yes | 14313 | 86.4 (85.2, 87.4) | < 0.001* | 7148 | 91.2 (89.8, 92.5) | < 0.001* | 7165 | 82.0 (80.3, 83.5) | < 0.001* |
| 6 | During pregnancy: blood sample taken |  |  |  |  |  |  |  |  |  |
|  | Yes | 14522 | 87.6 (86.6, 88.6) | < 0.001* | 7215 | 92.1 (90.6, 93.3) | < 0.001* | 7307 | 83.6 (82.1, 85.0) | < 0.001* |

ANC: antenatal care N<sup>a</sup>: weighted sample, %<sup>b</sup>: weighted percentage, S/N: serial number, <sup>c</sup>P-value for the analysis assessing yes vs no, \* Significant at 1% level.

### Factors associated with eight or more ANC contacts in Nigeria by rural and urban residences

Following adjustment for other predictors in the multivariable analyses, several factors were independently associated with ANC utilisation (of ≥ 8 contacts) in the overall Nigerian population (Table 3). These factors include the region of residence, rural-urban residences, maternal educational level, husband educational level, health insurance coverage, maternal age, maternal working status, wealth index, desire for pregnancy, birth order, maternal healthcare decision-making, maternal religion, and ethnicity (Table 3). The frequency of internet use was only marginally significant in the overall Nigerian population.

**Table 3:** Factors associated with ANC utilisation of ≥ 8 contacts in Nigeria across rural and urban residences.

| Factors | Overall (Nigeria) |  |  | Urban Nigeria |  |  | Rural Nigeria |  |  |
| --- | --- | --- | --- | --- | --- | --- | --- | --- | --- |
|  | AOR | 95%CI | P | AOR | 95%CI | P | AOR | 95%CI | P |
| **Region** |  |  | < 0.001* |  |  | < 0.001* |  |  | < 0.001* |
| North-Central | 2.42 | 1.79, 3.27 | < 0.001* | 7.44 | 4.64, 11.94 | < 0.001* | 1.29 | 0.87, 1.93 | 0.208 |
| North-West | 1.15 | 0.82, 1.63 | 0.416 | 3.39 | 2.00, 5.75 | < 0.001* | 0.68 | 0.44, 1.04 | 0.077 |
| South-East | 3.32 | 2.18, 5.07 | < 0.001* | 7.05 | 3.96, 12.57 | < 0.001* | 3.04 | 1.64, 5.64 | < 0.001* |
| South-South | 6.73 | 4.80, 9.44 | < 0.001* | 22.51 | 13.83, 36.63 | < 0.001* | 3.12 | 1.95, 4.99 | < 0.001* |
| South-West | 10.93 | 7.66, 15.60 | < 0.001* | 31.09 | 18.71, 51.67 | < 0.001* | 5.05 | 2.86, 8.91 | < 0.001* |
| North-East (Ref) | 1.00 | Reference | — | 1.00 | Reference | — | 1.00 | Reference | — |
| **Urban-rural residence** |  |  | < 0.001* |  |  |  |  |  |  |
| Urban | 1.33 | 1.17, 1.52 | < 0.001* |  |  |  |  |  |  |
| Rural | 1.00 | Reference | — |  |  |  |  |  |  |
| **Maternal education level** |  |  | < 0.001* |  |  | < 0.001* |  |  | < 0.001* |
| Primary | 1.40 | 1.13, 1.73 | 0.011** | 1.31 | 0.94, 1.84 | 0.111 | 1.47 | 1.13, 1.92 | 0.004* |
| Secondary | 1.69 | 1.37, 2.09 | < 0.001* | 1.65 | 1.19, 2.29 | 0.003* | 1.73 | 1.32, 2.27 | < 0.001* |
| Higher | 2.16 | 1.64, 2.84 | < 0.001* | 2.28 | 1.53, 3.38 | < 0.001* | 2.18 | 1.53, 3.11 | < 0.001* |
| No education | 1.00 | Reference | — | 1.00 | Reference | — | 1.00 | Reference | — |
| **Husband education level** |  |  | < 0.001* |  |  | 0.003* |  |  | 0.013** |
| Primary | 1.17 | 0.91, 1.50 | 0.219 | 1.11 | 0.73, 1.68 | 0.633 | 1.17 | 0.87, 1.57 | 0.291 |
| Secondary | 1.42 | 1.12, 1.80 | 0.004* | 1.34 | 0.89, 2.01 | 0.159 | 1.44 | 1.09, 1.89 | 0.01** |
| Higher | 1.85 | 1.39, 2.45 | < 0.001* | 1.85 | 1.15, 2.97 | 0.011** | 1.68 | 1.21, 2.34 | 0.001* |
| No education | 1.00 | Reference | — | 1.00 | Reference | — | 1.00 | Reference | — |
| **Health insurance coverage** |  |  | 0.001* |  |  | 0.043** |  |  | 0.014** |
| Yes | 1.64 | 1.22, 2.22 | 0.001* | 1.44 | 1.01, 2.05 | 0.043** | 2.09 | 1.16, 3.74 | 0.014** |
| No | 1.00 | Reference | — | 1.00 | Reference | — | 1.00 | Reference | — |
| **Maternal age** |  |  | 0.007* |  |  | 0.009* |  |  | 0.003* |
| 20-24 | 1.03 | 0.74, 1.43 | 0.85 | 1.21 | 0.70, 1.10 | 0.703 | 1.16 | 0.74, 1.82 | 0.515 |
| 25-29 | 1.17 | 0.84, 1.65 | 0.36 | 1.64 | 1.01, 2.65 | 0.048** | 1.11 | 0.71, 1.75 | 0.649 |
| 30-34 | 1.40 | 1.00, 1.97 | 0.05** | 1.75 | 1.06, 2.88 | 0.028** | 1.60 | 1.02, 2.53 | 0.042** |
| 35-39 | 1.46 | 1.02, 2.09 | 0.04** | 1.88 | 1.16, 3.03 | 0.01** | 1.53 | 0.95, 2.47 | 0.081 |
| 40-44 | 1.64 | 1.10, 2.43 | 0.015** | 2.15 | 1.27, 3.62 | 0.004* | 1.61 | 0.95, 2.73 | 0.075 |
| 45-49 | 1.31 | 0.81, 2.12 | 0.267 | 1.19<sup>d</sup> | 0.57, 2.49 | 0.572 | 2.09 | 1.14, 3.84 | 0.017** |
| 15 – 19 | 1.00 | Reference | — | 1.00<sup>e</sup> | Reference | — | 1.00 | Reference | — |
| Maternal working status |  |  | 0.007* |  |  |  |  |  | 0.011** |
| Yes | 1.20 | 1.05, 1.37 | 0.007* |  |  |  | 1.29 | 1.06, 1.56 | 0.011** |
| No | 1.00 | Reference | — |  |  |  | 1.00 | Reference | — |
| Wealth index |  |  | < 0.001* |  |  |  |  |  | < 0.001* |
| Rich | 1.47 | 1.22, 1.76 | 0.003* |  |  |  | 1.55 | 1.22, 1.98 | <0.001* |
| Middle | 1.29 | 1.09, 1.53 | < 0.001* |  |  |  | 1.45 | 1.19, 1.76 | <0.001* |
| Poor | 1.00 | Reference | — |  |  |  | 1.00 | Reference | — |
| Desire for pregnancy |  |  | 0.01 |  |  |  |  |  |  |
| Then vs. No more | 1.43 | 1.07, 1.92 | 0.017 |  |  |  |  |  |  |
| Later vs. No more | 1.24 | 0.87, 1.75 | 0.237 |  |  |  |  |  |  |
| No more | 1.00 | Reference | — |  |  |  |  |  |  |
| Birth order |  |  | < 0.001* |  |  | 0.001* |  |  | 0.001* |
| 1 | 1.45 | 1.22, 1.74 | < 0.001* | 1.44 | 1.14, 1.82 | 0.002* | 1.67 | 1.28, 2.18 | <0.001* |
| 2nd - 3<sup>rd</sup> | 1.28 | 1.13, 1.45 | < 0.001* | 1.349 | 1.15, 1.59 | < 0.001* | 1.30 | 1.08, 1.56 | 0.006* |
| ≥ 4 | 1.00 | Reference | — | 1.00 | Reference | — | 1.00 | Reference | — |
| Maternal healthcare decision |  |  | < 0.001* |  |  | < 0.001* |  |  | 0.029** |
| Respondent alone | 1.64 | 1.32, 2.02 | < 0.001* | 1.83 | 1.40, 2.41 | < 0.001* | 1.53<sup>b</sup> | 1.11, 2.12 | 0.01** |
| Respondent and spouse | 0.88 | 0.76, 1.02 | 0.081 | 0.874 | 0.72, 1.06 | 0.178 | 1.11<sup>a</sup> | 0.90, 1.36 | 0.332 |
| Spouse alone/someone else/others | 1.00 | Reference | — | 1.00 | Reference | — | 1.00<sup>c</sup> | Reference | — |
| Maternal religion |  |  | 0.006* |  |  |  |  |  | 0.002* |
| Christianity | 5.95 | 1.95, 18.17 | 0.002* |  |  |  | 9.50 | 2.77, 32.61 | < 0.001* |
| Islam | 5.42 | 1.75, 16.74 | 0.003* |  |  |  | 8.47 | 2.38, 30.14 | 0.001* |
| Traditionalist/others | 1.00 | Reference | — |  |  |  | 1.00 | Reference | — |
| **Ethnicity** |  |  | < 0.001* |  |  | < 0.001* |  |  | 0.002* |
| **Yoruba** | 1.93 | 1.32, 2.81 | < 0.001* | 2.07 | 1.31, 3.25 | 0.002* | 2.40 | 1.22, 4.73 | 0.011** |
| **Igbo** | 2.37 | 1.63, 3.45 | < 0.001* | 3.16 | 2.06, 4.85 | < 0.001* | 1.79 | 0.97, 3.31 | 0.062 |
| **Others** | 1.16 | 0.86, 1.58 | 0.33 | 1.53 | 1.05, 2.21 | 0.026 | 1.04 | 0.66, 1.65 | 0.870 |
| **Hausa/Fulani** | 1.00 | Reference | — | 1.00 | Reference | — | 1.00 | Reference | — |
| **Frequency of Internet use** |  |  | 0.049** |  |  |  |  |  |  |
| **< Once a week** | 0.84 | 0.60, 1.18 | 0.315 |  |  |  |  |  |  |
| **≥ Once a week** | 1.22 | 1.003, 1.48 | 0.047** |  |  |  |  |  |  |
| **Not at all** | 1.00 | Reference | — |  |  |  |  |  |  |
| **Frequency of watching TV** |  |  |  |  |  | 0.048** |  |  |  |
| **Not at all** |  |  |  | 1.32 | 1.04, 1.67 | 0.022** |  |  |  |
| **≥ Once a week** |  |  |  | 1.29 | 0.99, 1.68 | 0.062 |  |  |  |
| **< Once a week** |  |  |  | 1.00 | Reference | — |  |  |  |
| **Frequency of listening to radio** |  |  |  |  |  |  |  |  | <0.001* |
| **< once a week vs. Not at all** |  |  |  |  |  |  | 1.51 | 1.23, 1.86 | <0.001* |
| **≥ once a week vs. Not at all** |  |  |  |  |  |  | 1.69 | 1.37, 2.09 | <0.001* |
| **Not at all** |  |  |  |  |  |  | 1.00 | Reference | — |
| **Birth type** |  |  |  |  |  |  |  |  | 0.007* |
| **Multiple vs. Single** |  |  |  |  |  |  | 1.74 | 1.16, 2.61 | 0.007* |
| **Single** |  |  |  |  |  |  | 1.00 | Reference | — |

AOR: Adjusted odds ratio, CI: confidence interval, \* Significant at 1% level, \*\* Significant at 5% level, <sup>a</sup> Spouse alone/someone else/others, <sup>b</sup> Respondent alone, <sup>c</sup> Respondent and spouse (the reference category for this variable was changed to the 'respondent and spouse' so can make a meaningful comparison in this instance in urban residence), <sup>d</sup> 15 – 19, and <sup>e</sup> 45-49 (the reference category in this case was '45-49', which enables meaningful comparison in urban residence). We analysed factors associated with ANC utilisation in Nigeria using multivariable binary logistic regression. We only included variables with statistical significance ( $p < 0.05$ ) and identified significant factors using backward elimination at a 5% significance level ( $p < 0.05$ ). We reported AOR, 95% CI, and p-values for the variables retained in the final model. We used the same analysis procedure for rural and urban populations and considered potential confounders. The significant factors differ across the residences and leave the space vacant where a variable is not significant.

In urban residences, all geopolitical zones had significantly higher odds of ANC utilisation than the North-East region, with the South-West and South-South having substantially greater odds than others (Table 3). Mothers with at least a secondary level education (particularly those with higher education) had increased odds of ANC utilisation compared to their uneducated counterparts. This finding was slightly different for ‘husband’s education level’, where only the higher level of attainment increased the odds of ANC use. Access to health insurance significantly increased the odds of ANC use. Moreover, the odds of ANC use were noticeably higher among mothers aged 25 – 44 years. Compared to the birth order of four or more, the odds of ANC were higher for lower birth orders. Mothers who could decide independently on healthcare matters had greater odds of ANC use. Notably, the odds of using ANC were greater for all ethnicities compared to the Hausa/Fulani. Watching television at least once a week similarly increased the odds of ANC use in urban Nigeria.

In rural residences, the odds of ANC utilisation were significantly higher only in the southern geopolitical zones of South-West, South-East, and South-South (Table 3). All the other regions (northern geopolitical zones) had no statistically significant difference in their odds of ANC use compared to the North-East. Acquiring at least a primary level of education significantly increased the odds of ANC use, with the ‘higher level’ having the greatest odds. Mothers whose husbands acquired at least a primary education level had increased odds of ANC utilisation with greater odds for ‘higher education’ compared to the ‘no education’ category. The odds of ANC use similarly increased for mothers with access to health insurance coverage compared to those without such access. The odds of ANC use were only significantly higher among mothers aged 30 – 34 and those aged 45 – 49. Mothers that were working had greater odds of ANC use than those who were not. Belonging to middle or wealthy households increased the odds of ANC use. Compared to the birth order of four or more, lower birth orders were associated with increased odds of ANC utilisation. Mothers who could decide on their own concerning their health had greater odds of ANC use. Mothers who identified as Christians or Muslims had increased odds of ANC use compared to those who identified as Traditional or other religions. Compared to the Hausa/Fulani ethnic group, only the Yoruba had significantly increased odds of ANC use. Mothers who listened to the radio (compared to their counterparts who did not) and those who gave birth to multiple foetuses (compared to singletons) had increased odds of ANC use.

### Comparing factors associated with eight or more ANC contacts in rural and urban residences

In urban and rural residences across Nigeria, ANC utilisation shares some common factors, particularly the influence of education, access to health insurance, and maternal autonomy. Educational attainment consistently emerges as a significant factor in both settings. Mothers with at least a secondary education, particularly those with higher education, demonstrated increased odds of ANC use in rural and urban areas. Access to health insurance coverage also increased the odds of ANC use in urban and rural settings. Maternal autonomy (making independent decisions regarding healthcare matters) is a shared factor positively associated with ANC utilisation in both settings. Despite these commonalities, differences exist between urban and rural residences.

In urban areas, geographical disparities were prominent, with all the regions having significantly higher odds of ANC use than the North-East. In rural areas, the regional disparity follows a different pattern, with higher odds of ANC observed only in the southern regions—all the northern regions had substantially lower odds of ANC use than their southern counterparts. In urban areas, secondary and higher maternal education levels increased the odds of ANC use. In rural areas, primary, secondary, and higher education levels increased the odds of ANC use. In urban residences, only a ‘higher husband’s education’ level increased the odds of ANC use, while secondary and higher husband’s education levels were significant in rural settings. Maternal age between 25 – 44 years increased the odds of ANC use in urban residences, while specific age ranges of 30 – 34 and 40 – 45 demonstrate increased odds in rural areas. In urban areas, all ethnic groups had higher odds of ANC use than the Hausa/Fulani. In rural areas, only the Yorubas had greater ANC odds. The frequency of watching television was significant only in urban areas. Conversely, maternal working status, wealth index, birth type, maternal religion, and frequency of listening to radio attained statistical significance only in rural residences.

## Discussion

We conducted a comprehensive analysis of the 2018 NDHS data to determine ANC use, the receipt of ANC components and factors associated with ≥ 8 ANC utilisation in Nigeria, prioritising disparities across rural and urban residences in the country. The proportion of mothers having the recommended ≥ 8 ANC contacts was 20.3%—about 1.70-fold higher for urban residents (35.5%) and about 2-fold lower for mothers in rural areas (10.4%) in Nigeria. We previously investigated the rural-urban differences in ANC underuse based on the focused model and by analysing the 2013 NDHS [13]. Current findings are primarily based on the new WHO recommendation of ≥ 8 ANC contacts and may not be directly comparable to the results of our previous study. An estimated 42.2% of mothers recorded less than four ANC contacts (underuse, based on the focused model) in Nigeria—54% in rural and 23.9% in urban settings. Compared to the 2013 data, these findings would suggest a marginal decrease in ANC underuse in Nigeria (46.5% in 2013 [13]) and rural residences (61.1% in 2013[13]) but not in urban areas (22.4% in 2013 [13]). However, considering the potential influence of the WHO’s recommendation for ≥ 8 ANC contacts, it may be argued that there is no improvement in ANC utilisation in Nigeria. Our current findings underscore low utilisation of the recommended ANC in Nigeria with significant disparities between rural and urban settings (P < 0.001).

Moving to the components of ANC received, 69.3% of mothers had iron tablets or syrup nationally. In contrast, about one in every five mothers in urban areas and two out of every five in rural areas received no iron supplements. Our finding for the national estimates (69.3%) compares with a Nigerian study (using the same dataset) [14] but is lower than the results of a previous study (based on the 2013 NDHS), where 90.8% of mothers reported receiving iron tablets or syrup [25]. Our results, thus, indicate a decline in the receipt of this vital ANC component in Nigeria, with greater vulnerability for rural mothers. Additionally, the proportion of mothers who received medications for intestinal parasites (16%) is low but similar to the reports of previous studies of about 20% [25] and 16.7% [14] coverage in the overall Nigerian context. Iron supplementation and treatment for intestinal parasites are critical ANC components aimed at reducing anaemia in pregnancy [1, 2]. Anaemia contributes to general health issues, including lethargy, weakness, and less ability for daily activities [26]. In pregnant women, anaemia has substantial consequences for poor maternal and perinatal outcomes, including preterm birth, low birth weight, severe postpartum hemorrhage, and fetal malformation [27, 28], substantiating the importance of ensuring adequate receipt of ANC interventions.

We examined the adjusted association of ≥ 8 ANC contacts with several independent variables at national, urban, and rural levels. Overall, rural-urban residence was a significant predictor of ANC utilisation—an impetus for data disaggregation along this geographic divide for insights into the within-population differences, supporting our approach. However, rural-urban differences in the utilisation of maternal healthcare services are not new concepts; there is a consistent trend in previous studies indicating a higher uptake of maternal healthcare services among women in urban areas [13, 21, 29, 30]. This observation was confirmed in our analysis, and mothers in urban areas had higher odds of ANC use than their rural counterparts. There are well-established reasons for this phenomenon, including the availability of improved transport systems and other infrastructures or social amenities in urban relative to rural areas. These factors, along with well-established health services (with low staff turnover and potential for managing workforce transition [31]) and access to information, health facilities, and resources, can facilitate easier access to healthcare services or facilities for urban dwellers.

Our study also reveals considerable regional differences in the odds of ANC utilisation in Nigeria. While the observed regional gaps vary between national, rural, and urban residences (showing specific between-population differences), the southern regions generally had greater odds of ANC use than their northern counterparts, especially the North-East. The Hausa-Fulani are more predominant in parts of northern Nigeria, and the result associating the ethnic group with low odds of ANC use is consistent with the findings for northern Nigeria. Studies highlighting regional differences or gaps between southern and northern Nigeria are not new [5, 13, 31, 32]. The observed differences may reflect regional variations in security challenges, educational development, cultural or religious practices, healthcare services access and socioeconomic circumstances. For example, security challenges and less educational development are more pronounced in parts of northern Nigeria (the North-East and the North-West in particular) which can negatively impact healthcare services utilisation. In contrast to urban areas, where all regions had higher odds than the North-East region, there was no statistical difference in the odds of ANC use between regions in rural northern Nigeria. This finding implies a disproportionately low ANC use in all rural regions of northern Nigeria, which is a cause for concern. Our study, thus, highlights the urgent need to address the notable disparity between rural and urban residences in northern Nigeria to improve ANC use among residents, generally, but with priority for the most vulnerable sub-populations.

Notwithstanding rural or urban residences, our study reveals that mothers and their husbands/partners with higher levels of education had greater odds of achieving the recommended ANC contacts. This finding agrees with our previous study [13] and several national studies in Timor-Leste [33], Nepal [29], and African countries [30], further demonstrating the link between educational attainment and ANC utilisation. The consistency of this finding across all residences in Nigeria may also underscore the SDGs’ emphasis on the need for equal access to education for girls, indicating that education (SDG 4) can contribute to realising other SDGs such as SDG 3.1, which aims to reduce the global maternal mortality ratio to less than 70 per 100,000 live births by 2030. Notably, mothers who had autonomy in making decisions about their health were more likely to adhere to the recommended ANC contacts. While a previous study using the same data reported a similar finding at the national level [34], we analysed the data at rural and urban levels and found consistent results in all residences, lending credence to the importance of this factor for ANC utilisation.

Disparities in health services use between the rich and the poor are not new in public health; however, understanding the magnitude of the gaps is crucial for necessary intervention. Our study reveals that mothers from wealthy households were more likely to report having ≥ 8 ANC contacts. However, the differences in urban areas were not statistically significant after controlling for other factors. The differences remained significant in rural areas, potentially echoing the pronounced health equity gaps between the rich and the poor and emphasising the critical role of socioeconomic disadvantage in accessing ANC services in rural Nigeria. Consistent with this position, our utilisation analysis starkly highlights the situation, with mothers from wealthy families having almost seven-fold higher ANC contacts (30.9%) than those from low-income families (4.4%), even when both groups were from rural areas. These results underscore the negative impacts of low socioeconomic circumstances in utilising ANC services, a likely reason that access to health insurance was a significant predictor. Previous studies have reported the vulnerability of women from low-income families, resulting in lesser use of maternal health services in the overall Nigerian population [13, 35]. Current findings, however, emphasise the need to prioritise rural mothers for improved ANC utilisation, and (based on our study) addressing socioeconomic inequalities and providing health insurance coverage are pivotal entry points in this respect.

Young mothers, especially teenagers (15-19 years), were less likely to utilise ANC in our study than their older counterparts. Even within the teenage group, rural mothers had approximately four times less ANC utilisation than their urban counterparts. Current findings reveal a substantial disparity in ANC utilisation among young mothers, which is even worse for those living in rural areas. The results underscore the vulnerability of teenage mothers, who are often underprivileged, less educated, economically constrained, and at greater risk of being married off at a young age to relieve the economic pressure on their families. Present findings are consistent with our previous research based on the 2013 NDHS [13] and a recent study investigating the association of age differences on the ‘non-use of ANC’ services in Nigeria [14]. Our results also agree with a recent Nepalese study [36], which found that women aged 15-19 years were less likely to be autonomous or able to exercise their sexual and reproductive rights.

Our study demonstrates that mothers at four or higher parity were less likely to achieve the recommended ANC contacts. A few reasons may explain these results, including the low socioeconomic background of mothers with high parity. Similarly, this category of mothers tends to assume having gained experience through their previous pregnancy and childbirth and, therefore, less appreciative of the importance of the recommended ANC contacts. Our findings are similar to those of previous studies in Nigeria [13], Ghana [37], and other Sub-Saharan African countries [30]. Consistent with our previous study [13], mothers with multiple fetuses had greater odds of ANC use only in rural areas, which may be connected with the potential obstetric complication associated with delivering multiple babies.

Lastly, media and internet exposure demonstrate a significant association with increased odds of ANC use in different residences in Nigeria—internet in the overall, TV in urban areas, and radio in rural areas. These findings have considerable implications. Firstly, they highlight the potential influence of media exposure on maternal healthcare-seeking behaviour. Internet exposure, which was (marginally) significant nationally, suggests that digital platforms may play a role in disseminating information about ANC services, thereby increasing awareness and utilisation. A recent study made a similar finding concerning internet access and Caesarean Section utilisation in urban Nigeria [31]. The United Nations advocates unrestricted internet access as a basic human right [38, 39].

Moreover, Internet connectivity and digital literacy are known as “super social determinants of health” due to their potential to impact other social determinants of health [40, 41]. However, the proportion of mothers not using the Internet is high in Nigeria (90.6%), particularly in rural residences (97.3%). Given its potential to positively impact socioeconomic well-being and its association with ANC use in the current study, ensuring equitable access to the Internet may be critical to enhancing healthcare services utilisation in Nigeria, for example, through access to information, telemedicine, and so on. Secondly, the distinction in media influence between urban and rural areas is noteworthy. In urban settings, the association is with TV, suggesting that targeted health campaigns through the medium could be effective in urban areas. Conversely, in rural areas, the association is with radio exposure, revealing the continued importance of traditional media, in potentially influencing maternal healthcare behaviours. Radio, being widely accessible, can play a crucial role in reaching rural populations and encouraging ANC utilisation.

There are notable strengths in our study. First, we used the recent WHO guidelines [1, 7] of ≥ 8 ANC as the recommended number of contacts, providing results with the potential for immediate policy implementation in Nigeria. To our knowledge, this is the first research using the new WHO guideline to examine ANC utilisation and associated factors at Nigeria’s national, rural, and urban levels. Second, we performed analyses at overall, urban, and rural levels in Nigeria, enhancing the much-needed insights into the within and between population differences in the country. Our study, thus, provides evidence-based information for policies aimed at bridging equity gaps in maternal healthcare service use across geographic divides in Nigeria. This study used a nationally representative sample; therefore, findings are generalisable to the Nigerian population. Some limitations, however, need to be considered in our study. Firstly, the study used data collected based on maternal recall of their ANC contacts. Recalling≥ 8 ANC contacts may be challenging for some participants, which can contribute to over or under-reporting the number of ANC contacts achieved. However, in general, mothers tend to recall key childbirth events well, and we do not expect this limitation to alter the conclusion of our findings. Secondly, the study does not include all the variables that may be important for maternal healthcare service use. For instance, access to health facilities, travel distance, and key cultural, economic, and contextual barriers to accessing ANC are not exclusively included in the 2018 NDHS. Future studies may include those additional factors, including some qualitative data, to explore ANC use in Nigeria further. Lastly, given that the data utilised in our study were based on a cross-sectional design, inferring causality between outcome and predictor variables is beyond the scope of this study.

## Conclusions

In this comprehensive analysis of the 2018 NDHS data, we investigate ≥8 ANC utilisation and components in Nigeria, focusing on disparities between rural and urban areas. Our findings reveal a substantially low ANC utilisation in Nigeria, with only 20.3% of mothers achieving the recommended ≥8 ANC contacts. Moreover, notable rural-urban disparities persist with observable urban advantage across the board (35.5% ANC use in urban and 10.4% in rural areas), highlighting a critical need for targeted interventions to bridge the gap. The North-East region had the lowest ANC use at 3.7% and 3.0% in the national and urban areas, respectively, while the North-West had the lowest in rural areas (2.7%). Although the proportion of mothers with less than four ANC contacts has marginally decreased since 2013, the influence of the new WHO guideline suggests that overall ANC utilisation may not have improved in the country. Exploring ANC components, our study points to a concerning decline in iron supplementation and treatment for intestinal worms, nationally, but particularly impacting rural mothers, underscoring the urgent need for targeted interventions in this regard. Factors such as maternal and husband education, health insurance, and maternal autonomy were positively associated with increased ANC odds, regardless of residence. However, there were differences, with all ethnic groups having higher ANC odds than the Hausa/Fulanis in urban areas, while only the Yorubas had greater odds in rural areas. Internet use was significant only in the national context, watching television only in urban settings, while maternal working status, wealth, birth type, religion, and listening to the radio were significant only in rural areas. Efforts to bridge the rural-urban gap, address regional disparities, and prioritise vulnerable populations are imperative for enhancing ANC utilisation and maternal and perinatal health outcomes in Nigeria. Noteworthy strengths of our study include adherence to recent WHO guidelines, providing a basis for immediate policy implementation, and a pioneering examination of ANC factors at national, rural, and urban levels in Nigeria.

## Data Availability

The data analysed in the present study is accessible on the DHS program repository at https://www.dhsprogram.com/data/available-datasets.cfm. Access and data usage permission can be obtained through an online request on the DHS program website (www.dhsprogram.com).

## Acknowledgements

We thank the DHS Program for granting us access to the NDHS dataset analysed in this study. We also thank the participating mothers who shared their time and information. Thanks to Edith Cowan University, Western Australia, for making research facilities available to the lead author.

## Ethical approval and consent to participate

This study was based on a secondary data analysis of existing completely anonymised national data, and ethical approval is not required. Permission to use the dataset was obtained from the DHS Program, USA.

## Authors contributions

EOA and VK conceived and designed this study. EOA carried out statistical analysis, VO extracted data, populated Tables, and prepared Figure. EOA and MIA contributed to interpreting the results, while YZ provided additional statistical insights. EOA, AA, MIA, AAP, VO, YZ and VK contributed to writing and/or reviewed the manuscript for intellectual content. All authors approved the final copy for submission.

